# Impact of surface decontamination and systemic antimicrobials for surgical treatment of peri-implantitis: A systematic review and meta-analysis of randomized clinical trials

**DOI:** 10.1101/2021.02.26.21252213

**Authors:** Giacomo Baima, Filippo Citterio, Federica Romano, Giulia Maria Mariani, Fabrizio Picollo, Nurcan Buduneli, Mario Aimetti

## Abstract

**Background:** Efficient control of infection is essential to achieve desired outcomes in the surgical treatment of peri-implantitis lesions, although methods employed are largely heterogeneous.

**Purpose:** To compare the impact of different decontamination protocols and adjunctive systemic antimicrobials on the outcomes of surgical treatment of peri-implantitis.

**Materials and methods:** Randomized clinical trials (RCTs) on surgical treatment of peri-implantitis were selected through an electronic search on Medline, Embase, Scopus, and Central databases. Only studies comparing two or more anti-infective strategies were included. Following data extraction, two different sets of meta-analyses were performed. Firstly, overall impact of different implant surface decontamination methods was assessed by comparing baseline values with outcomes at 6-12 months. Secondly, pairwise comparisons evaluated the potential benefit of adjunctive systemic antimicrobials over placebo. Results were expressed as weighted mean effect (WME), weighed mean difference (WMD) or risk ratio (RR).

**Results:** Sixteen RCTs were included. No pairwise comparisons were available for different surface decontamination methods. Use of curettes resulted in improved probing depth (PD) (WME = 2.13 mm), but the results in terms of marginal bone levels (MBL) and percentage of disease resolution were unsatisfactory. Moreover, the adjunctive benefit of systemic antimicrobials over placebo was evaluated in two studies, representing a total of 178 implants. Despite not being statistically significant, the meta-analyses identified a higher probability of disease resolution (RR = 1.50) for test procedures. In terms of overall outcome, systemic antimicrobials with open flap debridement resulted in improved MBL (WME = 0.44 mm), reduced PD (WME = 2.46 mm) and 51.4% of disease resolution.

**Conclusions:** There is not enough evidence to support adjunctive usage of systemic antimicrobials together with the surgical treatment of peri-implantitis. Moreover, higher consistency is required to prove the superiority of a surface decontamination protocol over another (PROSPERO CRD42020182303).

**SUMMARY BOX:** *What is known:*

- Peri-implantitis is a common biological complication occurring at dental implants, and surgery is usually required to obtain thorough peri-implant infection control.
- No systematic reviews with meta-analysis have assessed surface decontamination protocols for surgical treatment of peri-implantitis, as well as the adjunctive benefit of peri-operative systemic antimicrobials.

*What this study adds:*

- This study offered the first evidence-based synthesis of randomized clinical trials regarding this relevant topic.
- Although protocol heterogeneity was high, a combination of mechanical and chemical implant surface decontamination is recommendable.
- Titanium brushes and local delivery of minocycline showed encouraging results; while the additional benefit of systemic antimicrobials needs to be further determined.

## 1. Introduction

Peri-implantitis is a rather frequent biological complication negatively affecting the success and survival of dental implants that is characterized by inflammation in the adjacent connective tissue and progressive resorption of supporting bone.^1^ Its estimated prevalence is around 22%, although there is a wide variation in case definitions and diagnostic criteria used across the studies.^2, 3^ Peri-implantitis has a bacterial etiology, and the success of treatment mostly depends on arresting the inflammatory process through efficient control of infection and removal of dysbiotic biofilm from the implant surface.^4^

Treatment of peri-implantitis traditionally involves a first phase of supramucosal plaque control and a subsequent deep debridement of the implant surface.^5^ Nonetheless, clinical success of non-surgical methods is limited in resolving peri-implantitis, and surgical intervention is usually recommended for better mechanical access to contaminated implant surfaces and efficient decontamination.^6, 7^ Although surgical procedures demonstrated favorable treatment outcomes in terms of reductions in probing depth (PD) and inflammation,^8^ complete disease resolution (DR) still remains unattainable for most cases.^9,10^

Insufficient implant decontamination may be regarded as a major explanation for these limited outcomes.^11^ Due to poor mechanical access to the bottom of bony defects and differences in micro and macro topography of titanium interfaces, appropriate surface decontamination continues to be a challenge.^12^ Therefore, there is an increasing interest on adjunctive agents that could possibly restore titanium biocompatibility and promote re-osseointegration.^13, 14^ Various chemical and mechanical methods including irrigation with saline, air powder abrasion, titanium brushes, citric acid application, ultrasonic and manual debridement, laser therapy, and topical medications have been proposed, but no single method was found to be superior in terms of clinical outcomes.^5, 15^ Furthermore, controversies still exist regarding the efficacy of adjunctive systemic antimicrobials in infection control and improvement of treatment outcomes.^9, 16, 17^

There are systematic reviews investigating the efficacy of surgical techniques and biomaterials in improving the clinical parameters of peri-implantitis lesions.^18, 19^ However, the clinical impact of available implant surface decontamination procedures has not been systematically evaluated so far. Proving the superiority of a specific method would be of uttermost relevance in enhancing the consistency of outcomes in peri-implantitis treatment and research. Accordingly, the evidence supporting usage of peri-surgical systemic antimicrobials is limited, and further data is required to justify their prescription. Therefore, the aims of the present systematic review were: 1) to evaluate and compare the efficacy of different implant surface decontamination protocols for the surgical treatment of peri-implantitis; 2) to investigate whether adjunctive systemic antimicrobials provide additional clinical benefits.

## 2. Materials and Methods

### 2.1 Report and protocol

This systematic review was reported according to the PRISMA statement^20^ and the protocol was registered on PROSPERO (CRD42020182303).

### 2.2 Focused questions

This systematic review was designed to answer the following focused questions:

*FQ1: In patients receiving surgical treatment of peri-implantitis, is any specific protocol of implant surface decontamination superior to others at improving the outcomes of therapy?*
*FQ2: In patients receiving surgical treatment of peri-implantitis, can adjunctive systemic antimicrobials provide any significant additional benefit in terms of clinical and radiographic outcomes?*

### 2.3 Eligibility criteria

Inclusion criteria were determined a priori and organized according to the PICOST acronym.

#### 2.3.1 PICO 1

(P) Population. Patients in good general health requiring surgical treatment of peri-implantitis.

(I) Intervention. Any type of chemical/physical or mechanical implant surface decontaminating agent, including possible combinations, used during surgical treatment of peri-implantitis.

(C) Comparison. Any possible comparisons between different protocols for intra-surgical decontamination.

(O) Outcome measures.

Primary outcome: changes in probing depth (PD) in mm.

Secondary outcomes: changes in radiographic marginal bone level (MBL), clinical attachment level (CAL), and bleeding/suppuration on probing (BoP/SUP). In addition, implant survival, disease resolution (DR) (possibly adhering to the definition to Carcuac et al.^21^ - residual PD ≤ 5 mm, no BoP, no suppuration, no progressive marginal bone loss after treatment), need of retreatment and patient-reported outcome measures (PROMs) were evaluated where available.

#### 2.3.2 PICO 2

(P) Population. Patients in good general health with a clinical diagnosis of peri-implantitis.

(I) Intervention. Surgical treatment of peri-implantitis together with adjunctive systemic antimicrobials.

(C) Comparison. Surgical treatment of peri-implantitis without adjunctive systemic antimicrobials.

(O) Outcome measures. See above.

#### 2.3.3 Studies and timing

(S) Types of studies. Randomized clinical trials (RCTs) on surgical treatment of peri-implantitis, with at least 6-month follow-up and a minimum of 10 patients (5 per group) were included. RCTs not directly comparing two anti-infective or surface decontamination agents or protocols were excluded.

(T). Timing. Data at 6-12 months after treatment were considered. If a study reported multiple evaluations between 6 and 12 months, data obtained at the latest time point were included. Data from longer follow-up periods were not used for the analysis, unless they were the only available.

### 2.4 Search methods for the identification of studies

The reviewing authors were calibrated before each phase of the study. Electronic search (Supplementary Table 1) was conducted on Medline (via PubMed), Embase, Scopus, and Cochrane Library electronic databases independently by two authors (GB and FC) with no restrictions on language, date of publication or publication status up to December 2020. In addition, hand searching (GB and NB) was performed on periodontics/implantology-related journals.

### 2.5 Study selection

Titles and abstracts (when available) of all identified studies were screened by two independent reviewers (NB and GB). Any disagreement was resolved by discussion with a third reviewer (FC). Full text of studies of possible relevance or for which there was insufficient data in the title and abstract were assessed independently by two reviewers (NB and FC). Differences between them were settled by a third reviewing author (GB). The reasons for exclusion after the full text analysis were recorded.

### 2.6 Data extraction and management

Data from included studies were extracted by two reviewers (GB and FC) independently using predefined data extraction forms (Table 1). If necessary, corresponding authors of the included studies were contacted for clarification of any missing information. If no reply was received within three months, the study was excluded.

**Table 1.**
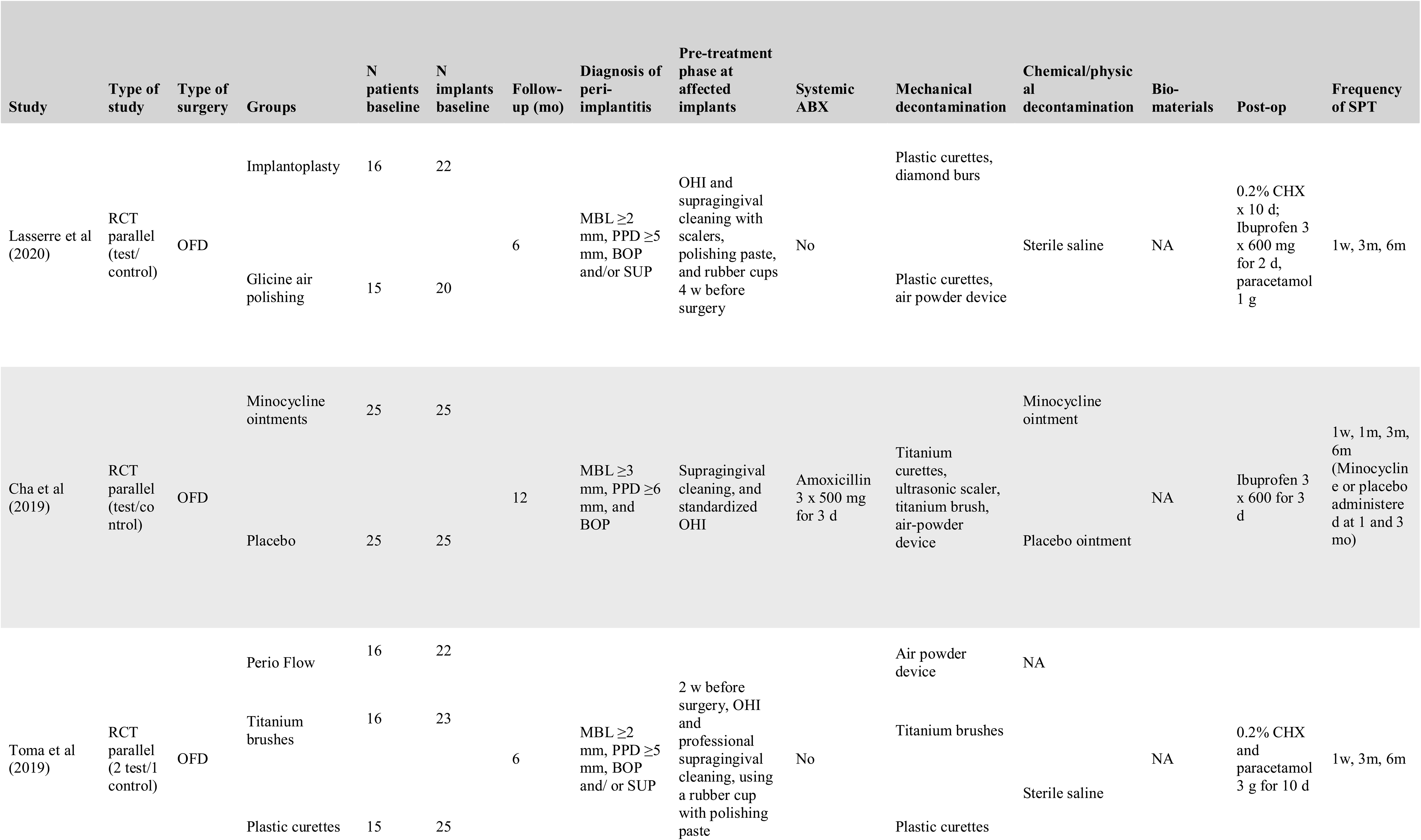
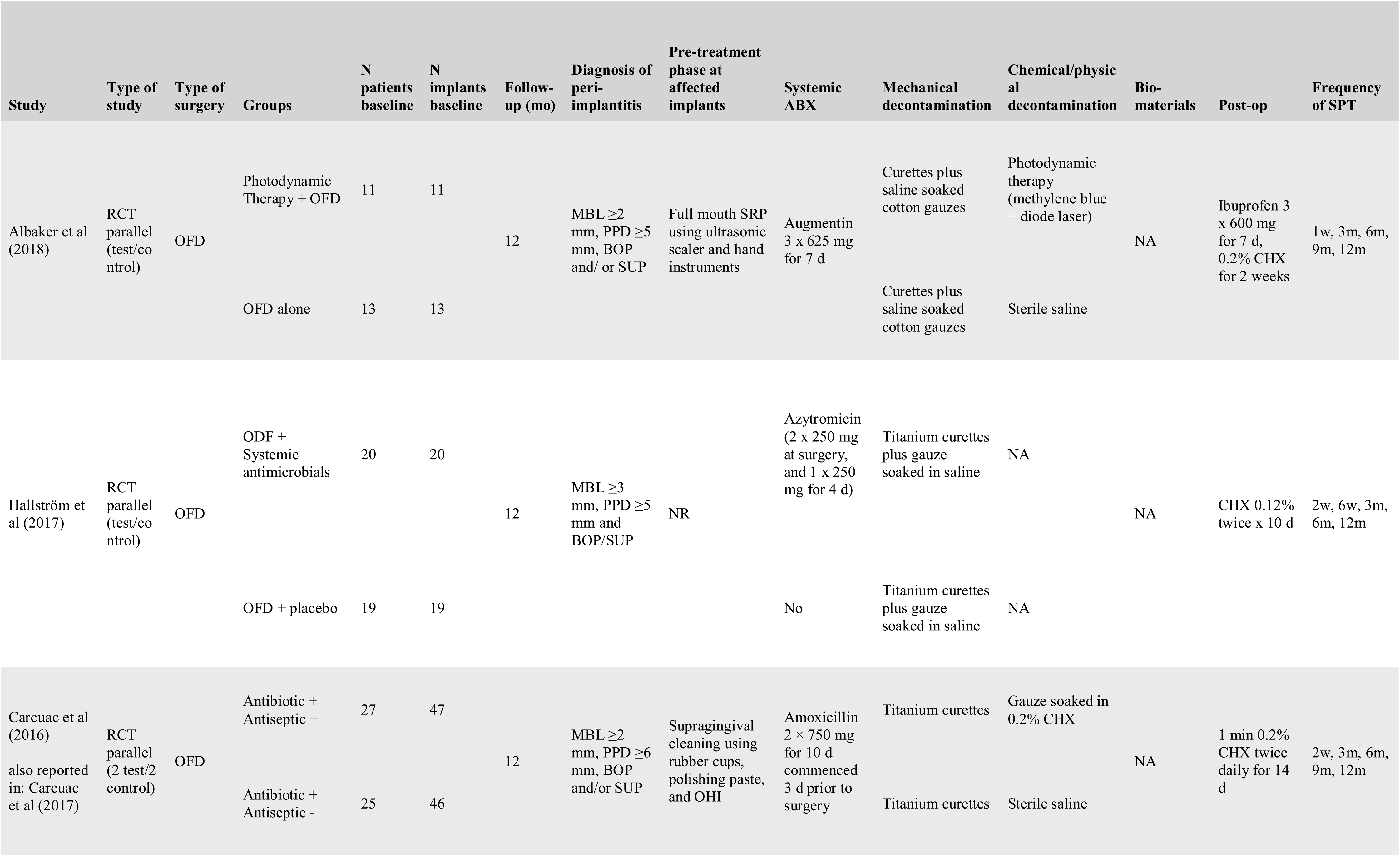
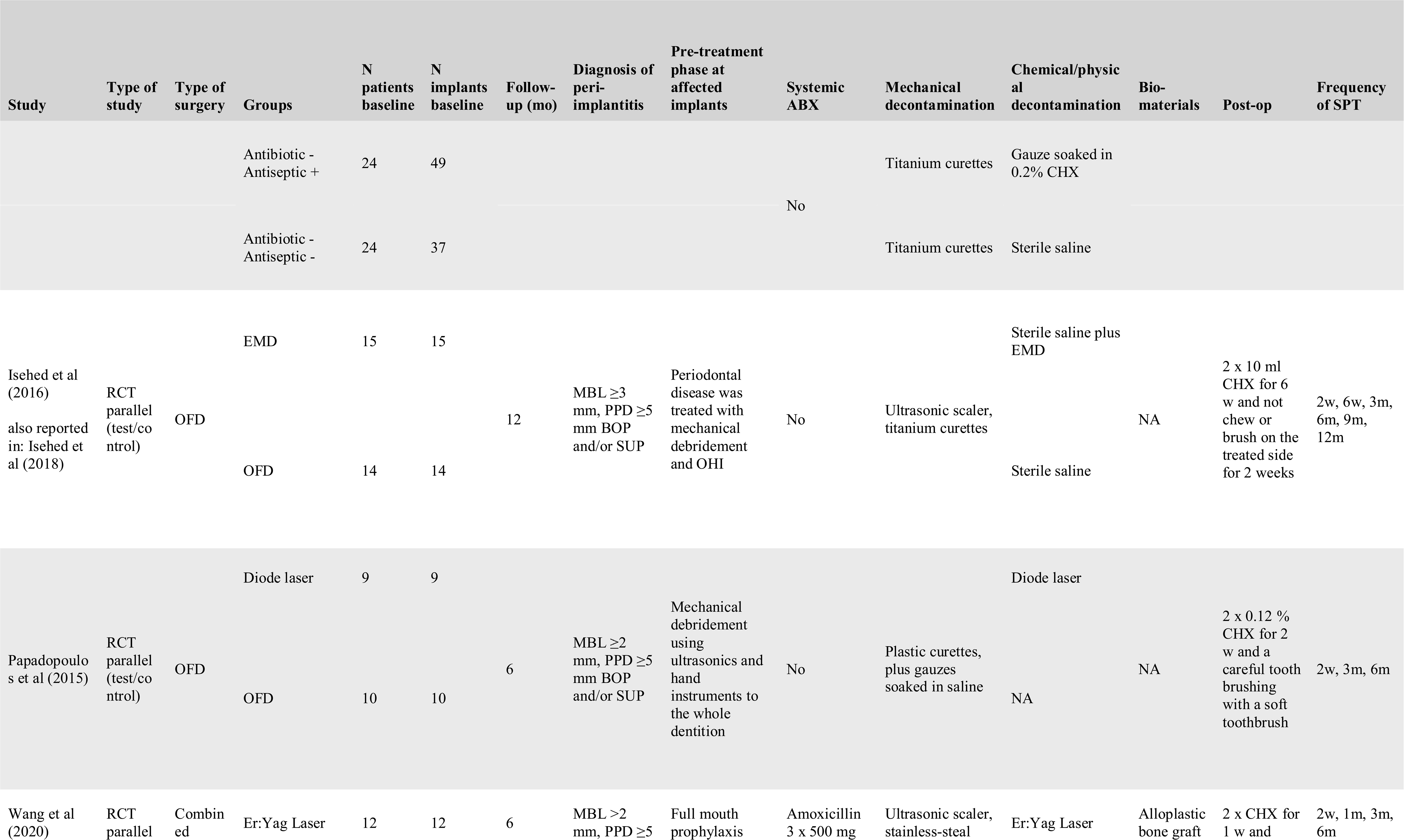
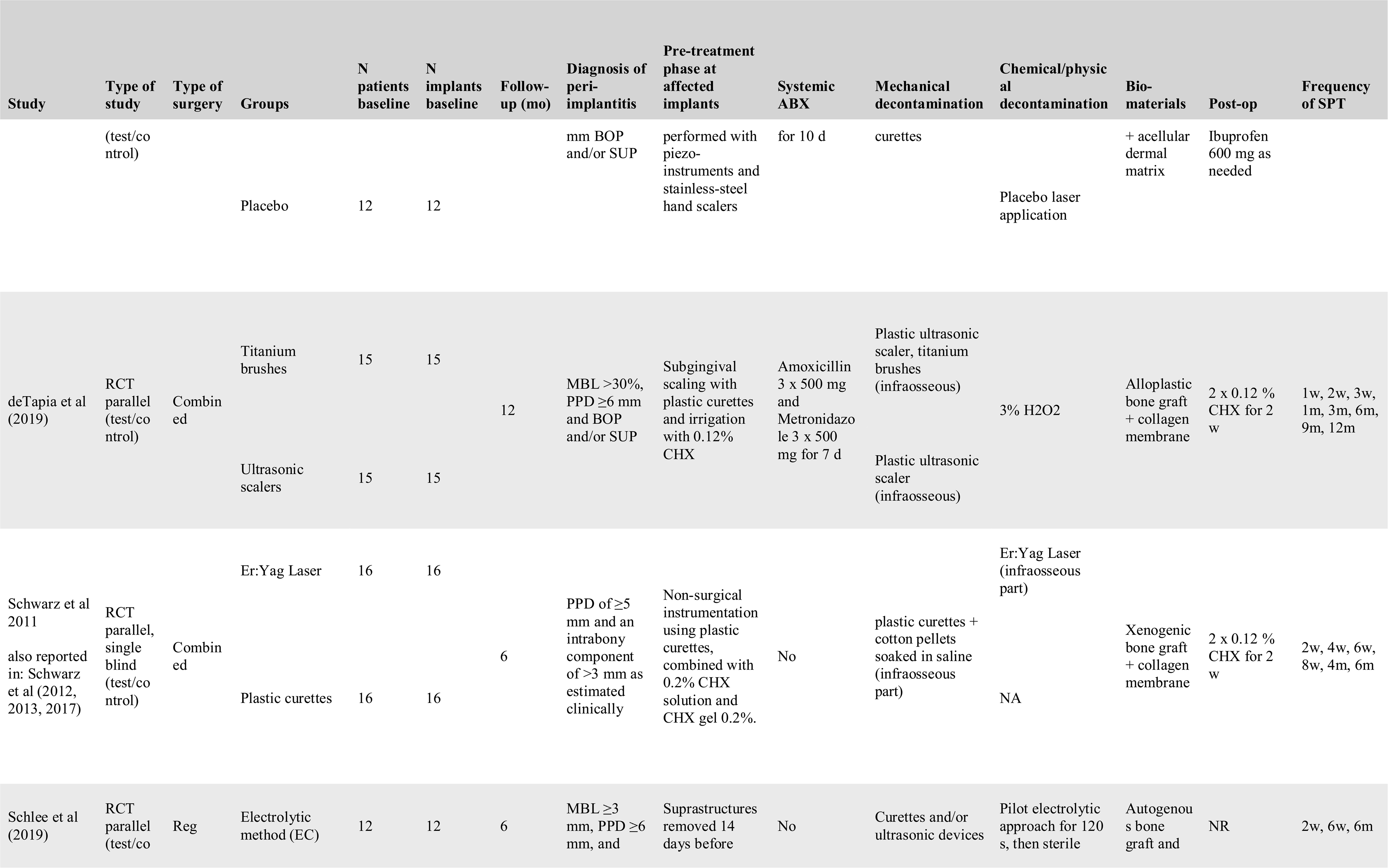
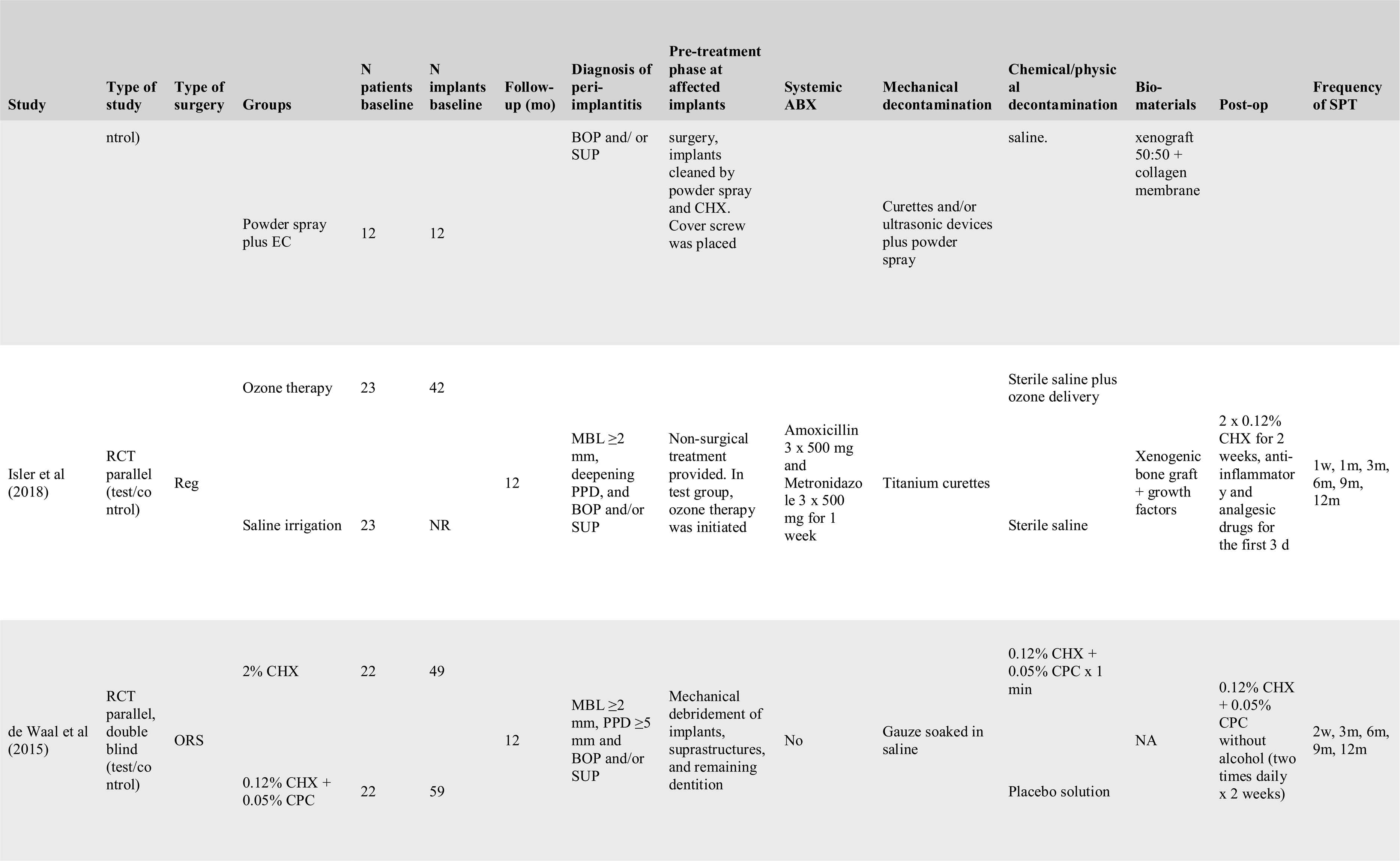
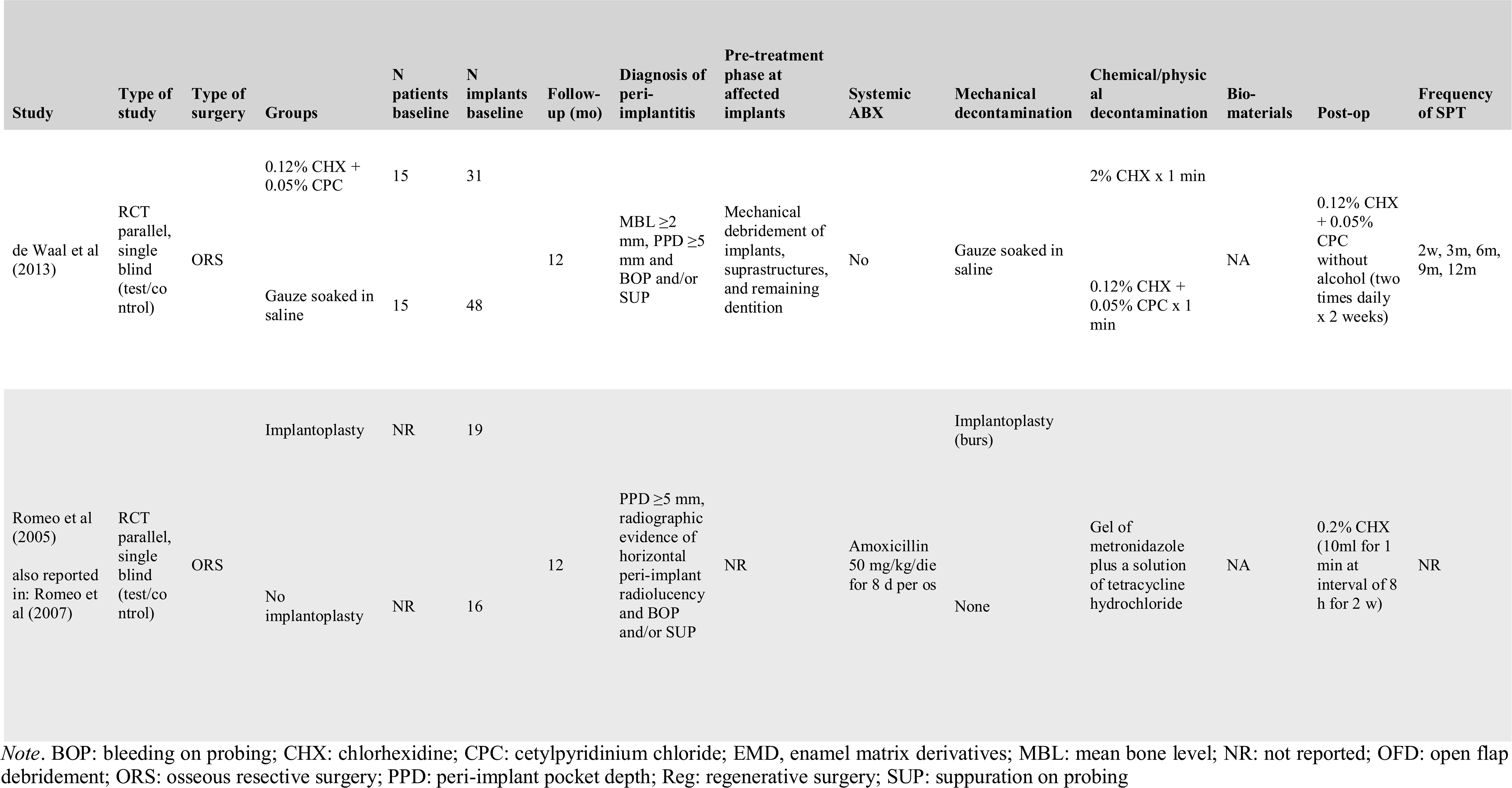
Included studies: anti-infective protocols employed

Data on general information (first author, year of publication and setting); methods (study design, diagnostic criteria for peri-implantitis, sample size, follow-up period, population); interventions and controls (pre-treatment phase, type of surgery, type/combination of decontaminating agents, biomaterials, post-surgical care) and outcomes (changes in radiographic MBL, CAL, PD, REC percentage of implants with BoP and/or suppuration, implant survival, DR, PROMs) were tabled.

### 2.7 Assessment of risk of bias in the included studies

The risk of bias in the included studies was assessed independently and in duplicate by the two reviewing authors (GB and FC) according to RoB.2 tool.^22^

### 2.8 Data analyses

For continuous outcomes at 6-12 months (changes of MBL and PD reduction), mean values and standard deviations were combined and analyzed with weighted mean effect (WME) and 95% confidence intervals (CIs). Dichotomous data were pooled as weighted mean percentage and 95% CIs. In pairwise comparison, the estimates of the effect were expressed as weighted mean differences (WMD) and 95% CIs for continuous outcomes and as risk ratio (RR) and 95% CIs for dichotomous outcomes. In order to account for within-patient correlation in studies which failed to adjust for it, an intracluster correlation coefficient of 0.07 was assumed for the calculation of the effective sample size and CIs. Study-specific estimates were pooled with and random-effects models to account for between studies heterogeneity.^23^. The Mantel–Haenszel method was performed to combine the dichotomous outcomes, and the inverse of variance method to combine the continuous outcomes.

Two different sets of analyses were conducted. First, the outcome of different protocols for the decontamination of the implant surface was assessed by comparing baseline values with outcomes obtained with follow-up longer than 6 months. Only protocols reported exactly the same procedures, including the type of surgery [i.e. open flap debridement (OFD), resective or regenerative], in two or more studies were included. For studies with multiple arms, each decontaminating intervention was considered separately. Secondly, RCTs were used to evaluate the potential benefit of adjunctive systemic antimicrobials in OFD. For this analysis, no discrimination was made between the different methods of surface decontamination. OFD without systemic antimicrobials was considered as control.

Statistical heterogeneity among studies was explored using the *I*^2^ index^24, 25^ and the Cochrane’s Q statistic (*p* < 0.1). For each meta-analysis forest plots were generated. Statistical significance was set to *p* < 0.05. Statistical analyses were performed using statistical software package OpenMeta [Analyst].^26^

## 3. Results

### 3.1 Search

As outlined in Supplementary Figure 1, the electronic search yielded 1,835 publications and hand searching identified 5 additional studies. After removal of duplicates, the total number of articles screened was 1,497. Twenty-five records were identified as potentially relevant during screening (Kappa = 0.63, substantial agreement). Of these, four records were excluded after full text reading (Kappa = 0.911, almost perfect agreement), while six records were excluded as double-data, with the most relevant data retained for analysis.^9, 27–31^ Finally, 16 RCTs were included in the qualitative analysis and 9 were used for meta-analyses. Reasons for article exclusion were reported in Supplementary Table 2.

### 3.2 Description of selected studies

#### 3.2.1 Design

As shown in Table 1, all RCTs had a parallel arm design, 14 studies included one experimental group and one control group and two studies adopted multiple interventional arms.^21, 32^ Six studies included multiple dental implants patient and did not account for clustering. Of these, five were included in the meta-analyses after correcting for the within-patient correlation.^21, 32–35^ Six studies reported results at 6 months, and 6 studies at 12 months.

#### 3.2.2 Study sample

Sample size varied from 17 to 100. All trials were carried out in European Universities, except from one that was performed at the Yonsei Dental Hospital of Seoul,^36^ one at the University of Michigan,^37^ and one in a private practice in Saudi Arabia.^38^ Diagnosis of peri-implantitis followed the last update of the World Workshop on the Classification of Periodontal and Peri-Implant Diseases^39^ only in two studies;^36, 40^ but was consistent across all studies except for minor differences (either MBL ≥ 2 or 3 mm together with PD ≥5 or 6 mm and BoP/SUP). Only three studies considered the intrasurgical morphology of the bony defect among inclusion criteria.^41–43^ Mean age of patients ranged from 54.3 to 71.7 years, while the proportion of females varied from 25% to 80.9%. In three studies smokers were excluded;^33, 36, 37^ two studies did not report any information on smoking habits;^32, 44^ while the remaining trials included a variable number of smokers, ranging from 14.2% to 50%. Five studies presented implants with machined surface, with proportions ranging from 1.3 to 35%; four studies did not treat turned implant surfaces,^32, 33, 36, 37^ without this being a specific exclusion criteria; while for seven studies this information was not available.

#### 3.2.3 Interventions

In total, 849 implants were treated in 604 patients. Eight studies performed a non-surgical deep implant debridement prior to the intended treatment; while the other eight only performed supragingival scaling or did not report this information.^17, 21, 32, 33, 36–38, 44^ Systemic antibiotics were administered prior to the surgical treatment in two RCTs;^21, 45^ in six studies they were prescribed on the day of the surgery;^17, 36–38, 42, 43^ while in the other eight studies no systemic antimicrobials were used. Eight studies reported decontaminating protocols applied to OFD;^17, 21, 32, 33, 36, 38, 44, 46^ five studies to reconstructive or combined procedures;^37, 40–43^ and three studies to apically positioned flap and osseous resective surgery (ORS).^34, 35, 45^ Five RCTs used mechanical decontamination,^17, 32, 33, 41, 44^ two studies chemical decontamination,^34, 35^ while nine studies a combination of both. Curettes were the most preferred method for mechanical debridement, used in 25 arms of 12 RCTs, either alone or in combination with other devices; together with gauzes soaked in saline in 16 arms of 7 studies. Ultrasonic scalers were adopted in 10 arms of five studies; powder spray in five arms of four studies; titanium brushes in four arms of three studies, while implantoplasty was carried out in six arms of four studies. Among chemical and physical decontaminations, chlorhexidine (CHX) at 0.12 or 0.2 formulations was applied in four arms of three studies; lasers were used in three test groups of three different studies; a gel of metronidazole followed by a solution of tetracycline hydrochloride was rubbed on implant surfaces in both arms of the study by Romeo et al.;^45^ while enamel matrix derivatives (EMD);^46^ ozone therapy;^42^ photodynamic therapy;^38^ electrolytic current;^40^ and minocycline ointments^36^ were part of only one study arm. The main outcomes of the included studies are presented in Table 2.

**Table 2.**
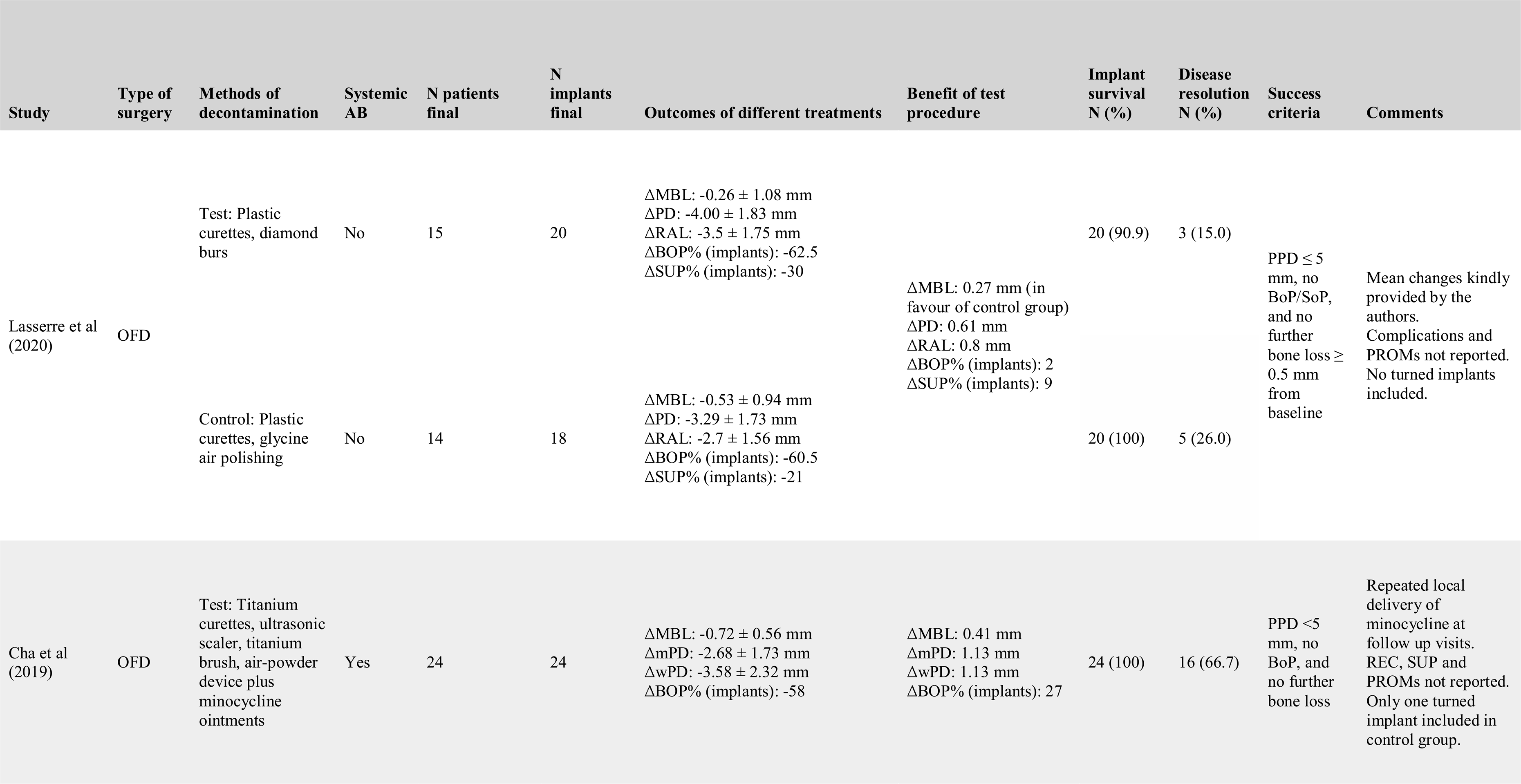
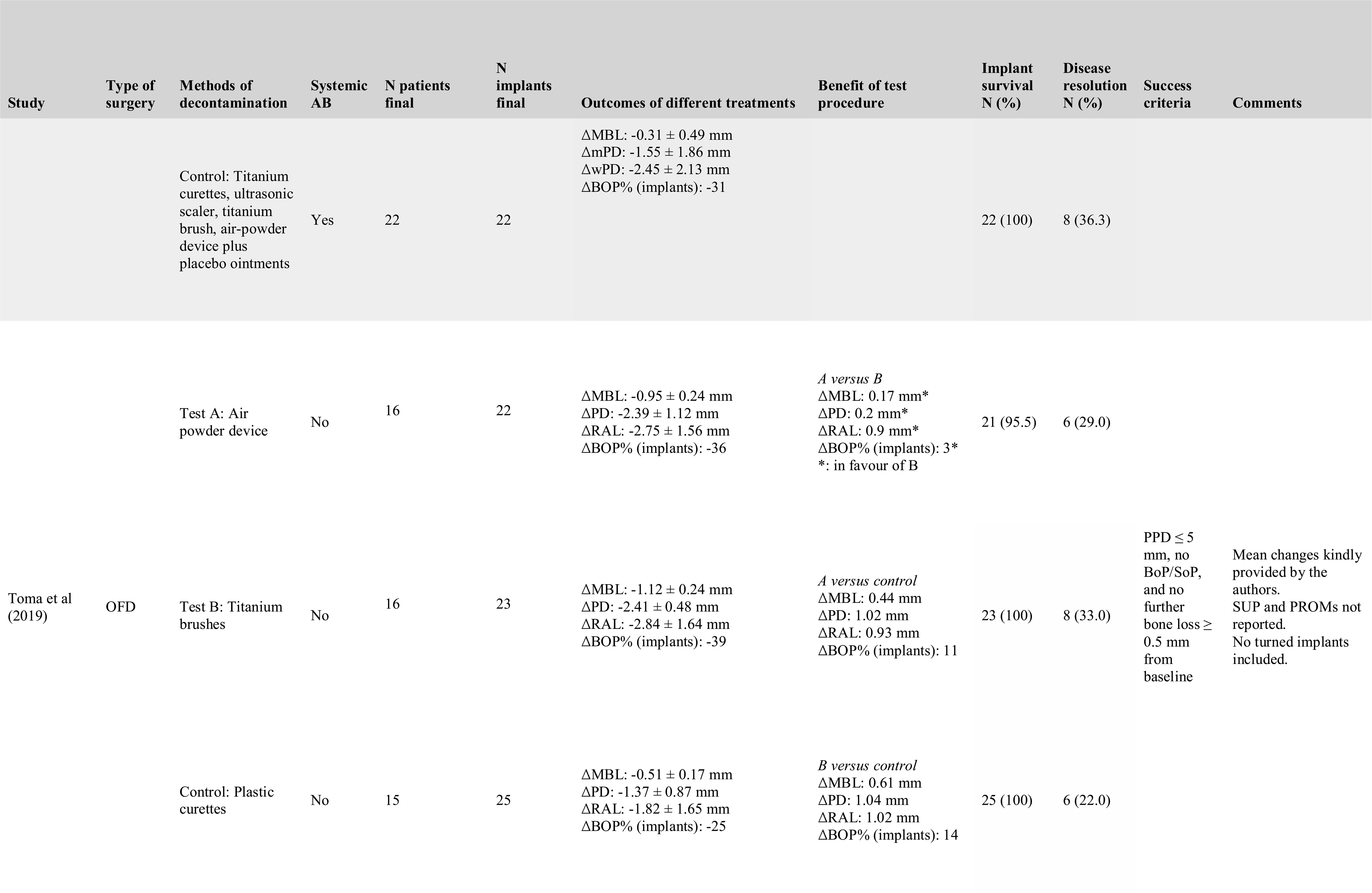
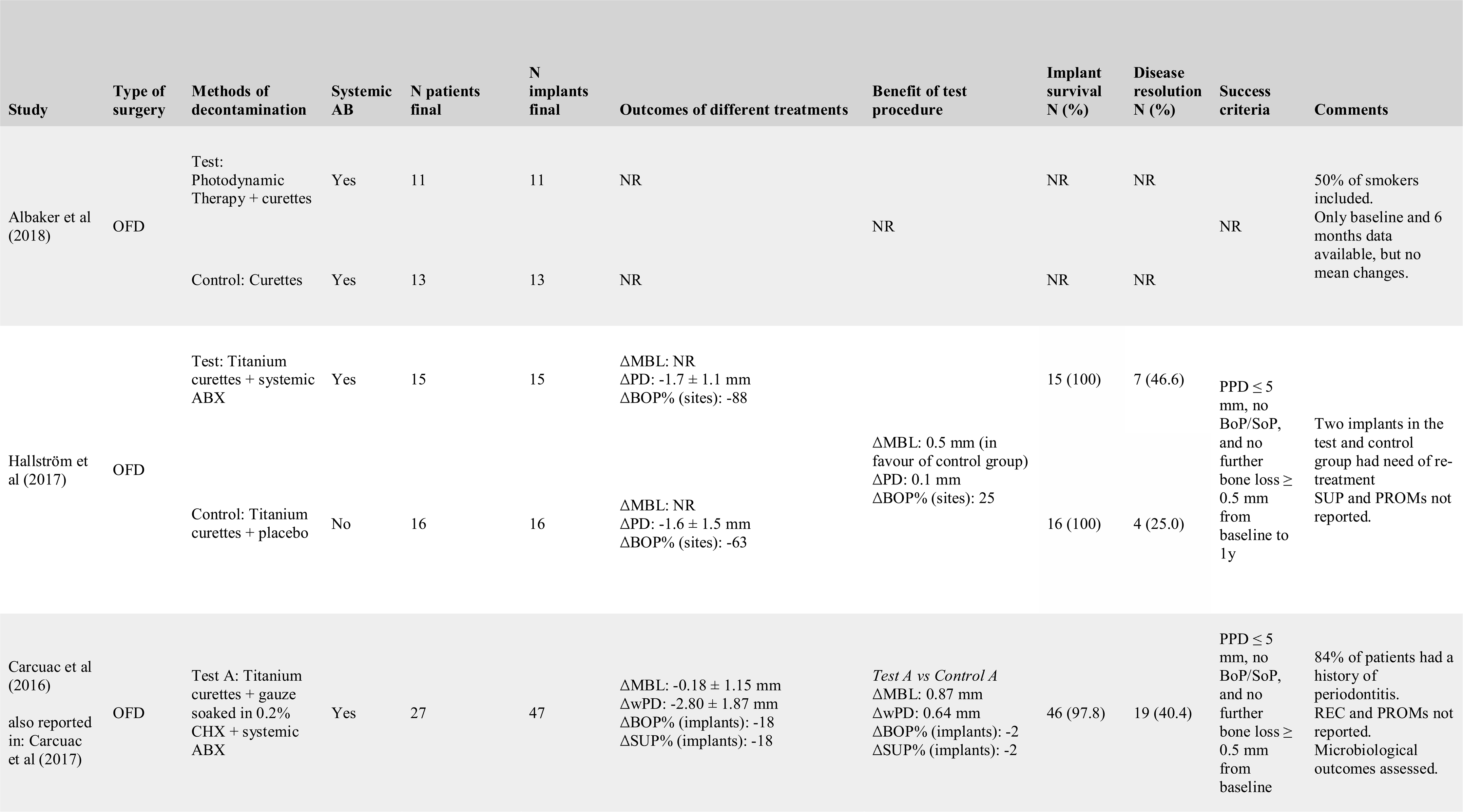
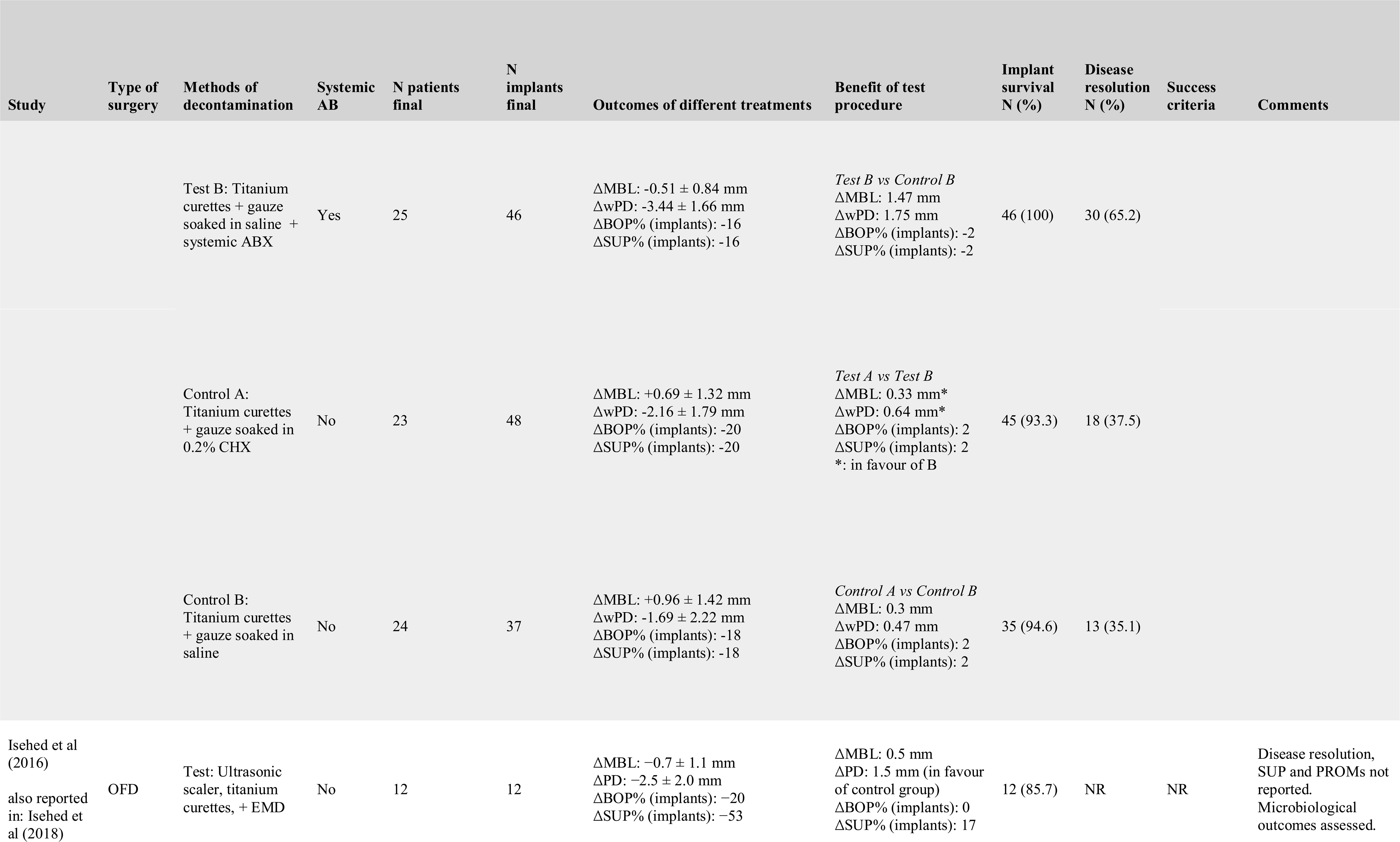
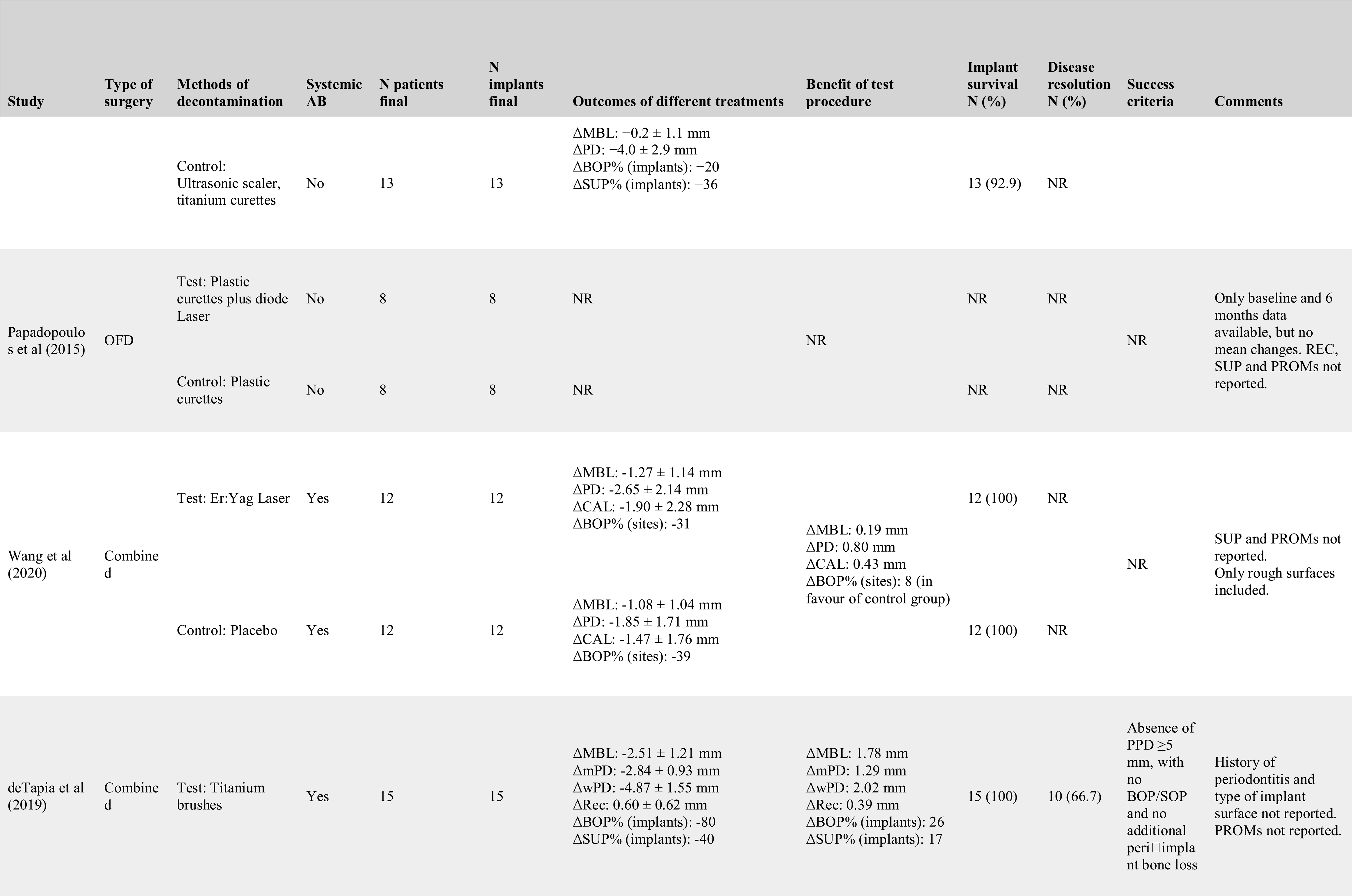
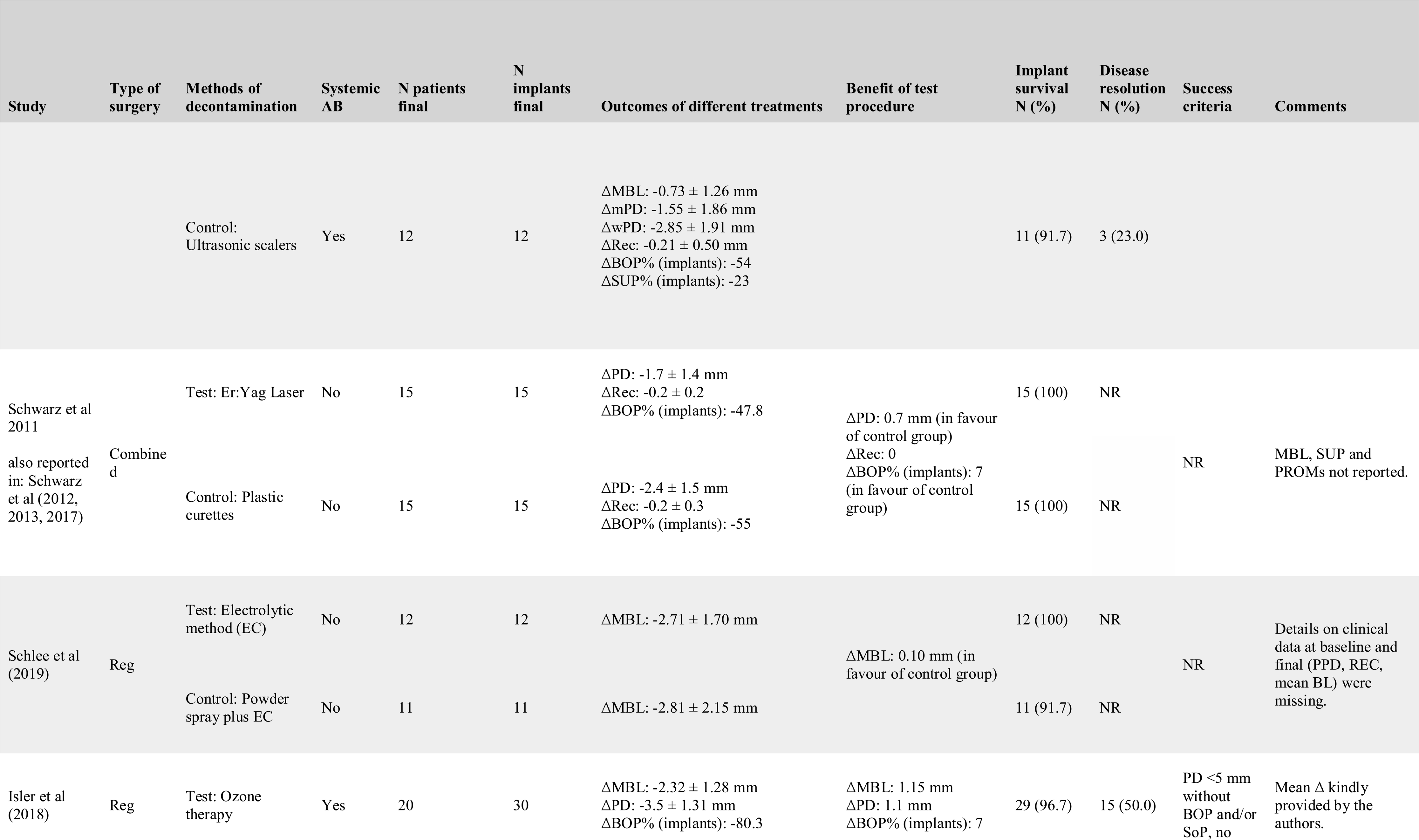
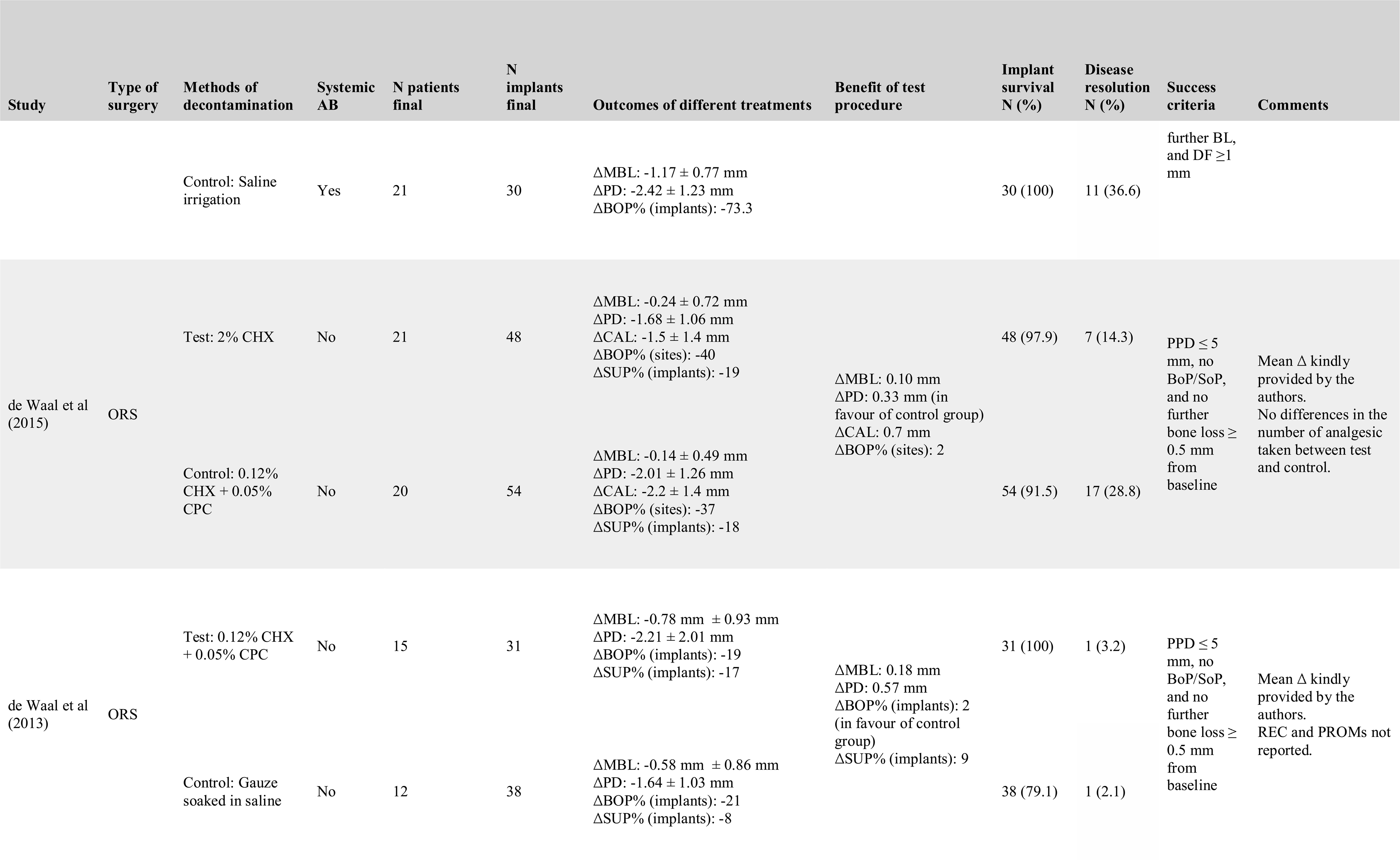
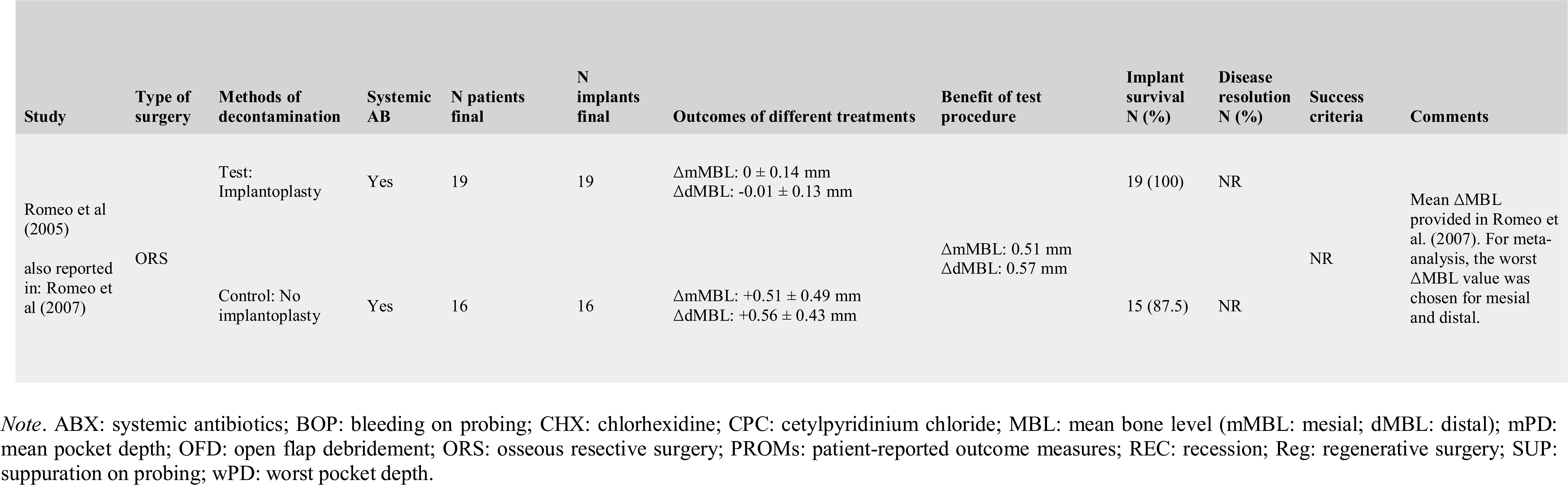
Included studies: summary of outcomes

### 3.3 Risk of bias in individual studies

The risk of bias assessment for the included RCTs is summarized in Figure 1. Ten papers had a low risk of bias. Three studies exhibited selection bias,^32, 33, 42^ one attrition bias,^38^ and one missing outcome data.^44^ One study was considered to have a high risk of bias due to the selection of reported results.^40^

**Figure 1.**
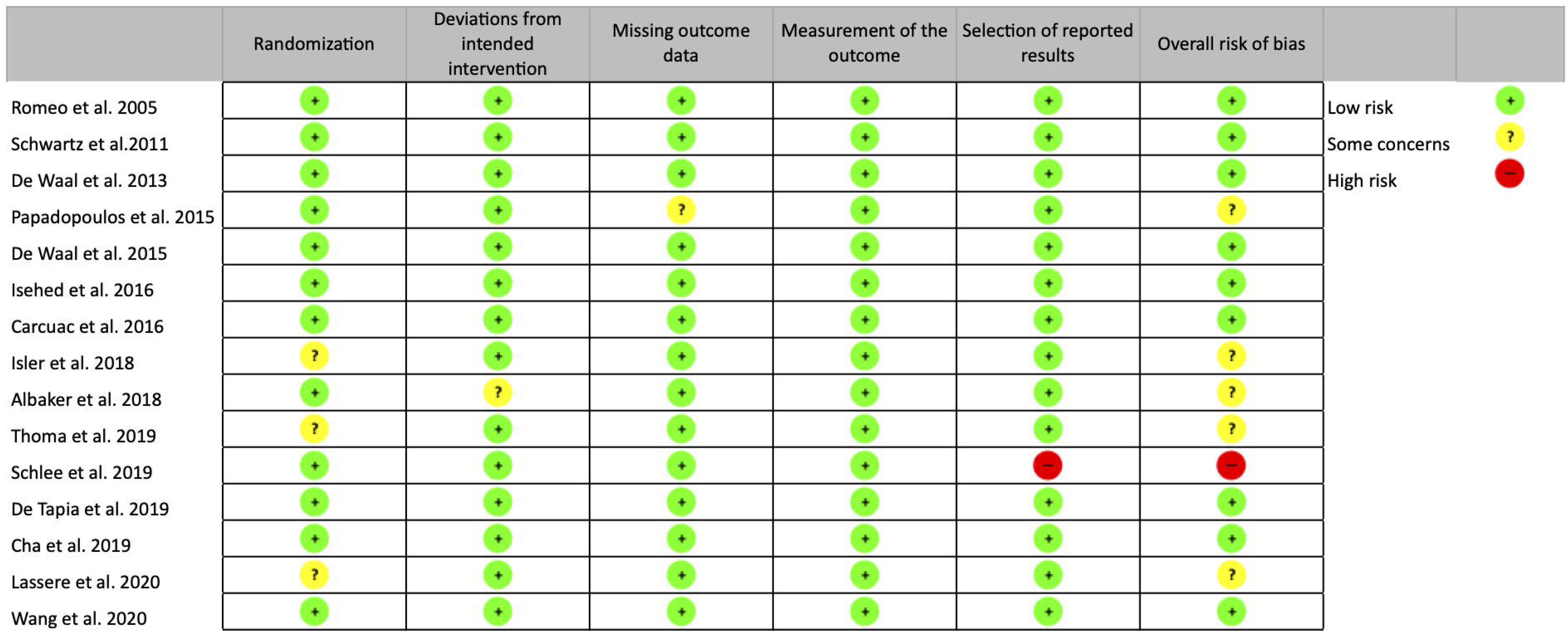
Risk of bias for studies in the systematic review.

### 3.4 Outcomes of different decontamination protocols in OFD

A total of 452 implants in 339 patients were treated by OFD procedures. The MBL change ranged from a reduction of - 0.96 mm in a control arm in which only titanium curettes plus gauze soaked in saline were employed,^21^ to a gain of 1.12 mm with the use of titanium brushes.^32^ The single use of curettes for surface decontamination resulted in a negligible MBL reduction (WME = −0.05 mm, 95% CI: −0.90/0.81) in three studies (three arms), with high heterogeneity (*I*^2^ = 94.82 p < 0.001). In terms of PD reduction, the analysis included four studies (4 arms) yielding a WME of 2.14 mm (95% CI: 1.07/3.20); while the weighed mean percentage of DR amounted to 25% (95% CI: 16/33) with no heterogeneity (Figure 2A,B,C).

**Figure 2.**
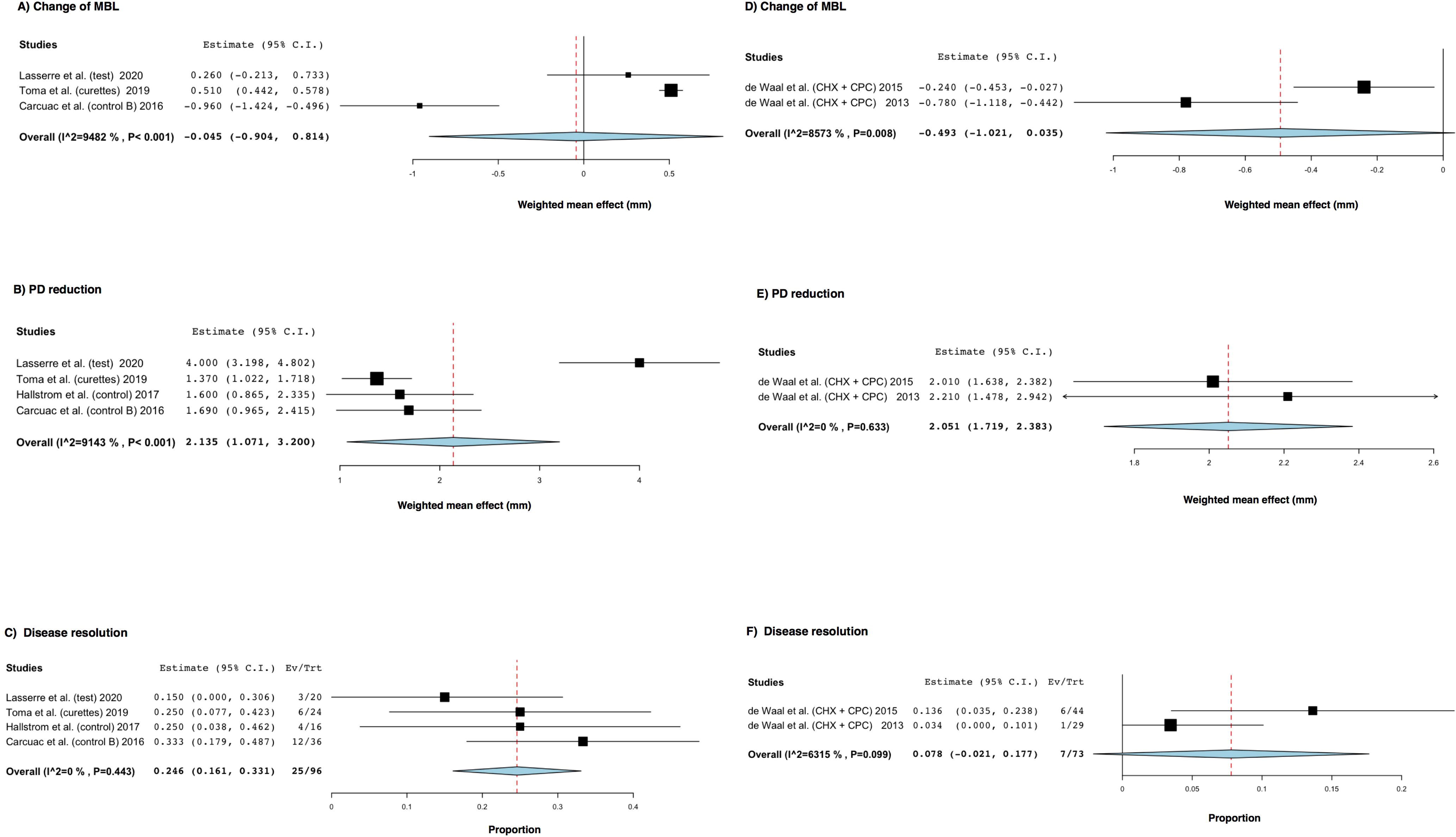
Forest plots of weighted mean effect (WME) for different clinical parameters after implant surface decontamination with curettes or chlorhexidine (CHX), and cetylpyridinium chloride (CPC) without systemic antimicrobials. A) Marginal bone levels (MBL) changes with curettes in open flap debridement (OFD), B) Pocket depth (PD) reduction with curettes in OFD, C) Disease resolution with curettes in OFD. D) MBL changes with CHX + CPC in osseous resective surgery (ORS), E) PD reduction with CHX + CPC in ORS, F) Disease resolution with CHX + CPC in ORS.

None of the studies gave information on defect fill or PROMs, and implant survival was described in only two with results ranging from 90.9% to 100% on the implant level.

### 3.5 Outcomes of different decontamination protocols in reconstructive surgery

A total of 192 implants in 156 patients were treated using reconstructive or combined procedures. None of the study arms provided comparable decontamination protocols, thus data meta-analysis was not possible.

### 3.6 Outcomes of different decontamination protocols in resective surgery

A total of 222 implants in 156 patients were treated by resective procedures. The use of CHX and cetylpyridinium chloride (CPC) together with ORS was investigated in two arms of two different studies by the same research group and no systemic antibiotics were used.^34, 35^ WME in terms of post-treatment MBL was −0.49 mm (95% CI: −1.02/0.04) with significant heterogeneity (*I*^2^= 85.73; p = 0.008), WME for PD reduction was 2.05 mm (95% CI: 1.722.38) with no heterogeneity (*I*^2^= 0; p = 0.633). Eighteen out of 85 implants were successfully treated, and this accounted for a weighted mean percentage of DR of 17% with significant heterogeneity (*I*^2^= 92.72; p < 0.001) (Figure 2D,E,F).

### 3.7 Potential advantages of adjunctive systemic antimicrobials

Only two studies directly compared OFD with and without adjunctive systemic antibiotics.^17, 21^, while no data were available for the other surgical treatment approaches. The analysis evaluated 178 implants for PD reduction as outcome and a total of implants ranging from 178 to 209 for the secondary outcomes. Implant survival rate was comprised between 93.3% and 100%.

Figure 3 depicts the results of the pairwise meta-analysis for MBL changes. Only one study with four arms reported on radiographic changes. A statistically significant benefit (WMD = 1.17 mm; 95% CI: 0.58/1.76, p<0.001) in terms of MBL changes was observed in favor of the adjunctive systemic antimicrobials at 12 months, with a heterogeneity of 60.72%.

**Figure 3.**
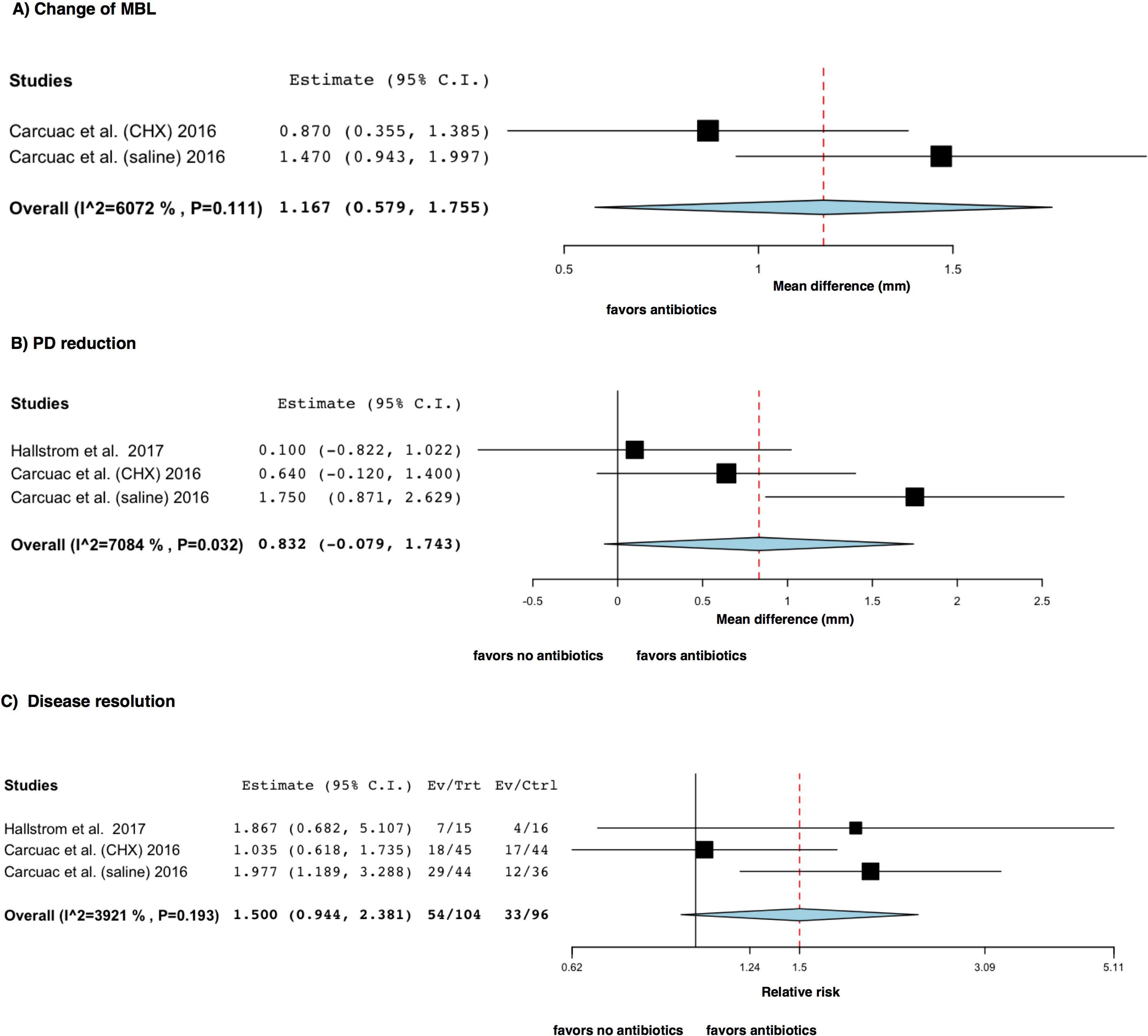
Forest plots of the pairwise meta-analyses of open flap debridement (OFD) with antibiotics vs. OFD without antibiotics for A) Marginal bone levels (MBL) changes (weighted mean difference, WMD), B) Pocket depth (PD) reduction (weighted mean difference, WMD), and C) Disease resolution (risk ratio, RR).

PD reduction was reported in two studies accounting for a total of six arms, failing to identify a statistically significant benefit at the test sites (WMD = 0.83 mm; 95% CI: −0.08/1.74). Disease resolution according to the criteria of Carcuac et al.^21^ was reported in the same two studies, with a RR of 1.50 (95% CI: 0.94/2.38; p = 0.086) favoring the antibiotics administration with low heterogeneity. None of the studies reported on defect fill or PROMs, while implant survival over one year or more was described in two.

Figure 4 illustrates changes in clinical parameters from baseline to 6-12 months postsurgery around implants treated using OFD with or without systemic antimicrobials. When adjunctive systemic antimicrobials were prescribed together with OFD (Figure 4A,B,C), MBL changes amounted to 0.44 mm (95% CI: 0.22/0.67) based on four arms in two studies. Regarding the secondary outcomes, PD reduction was assessed in five arms of three studies and amounted to 2.46 mm (95% CI: 1.74/3.18); while mean percentage of DR was 51% (95% CI: 33/69), and was based on three arms of two studies.

**Figure 4.**
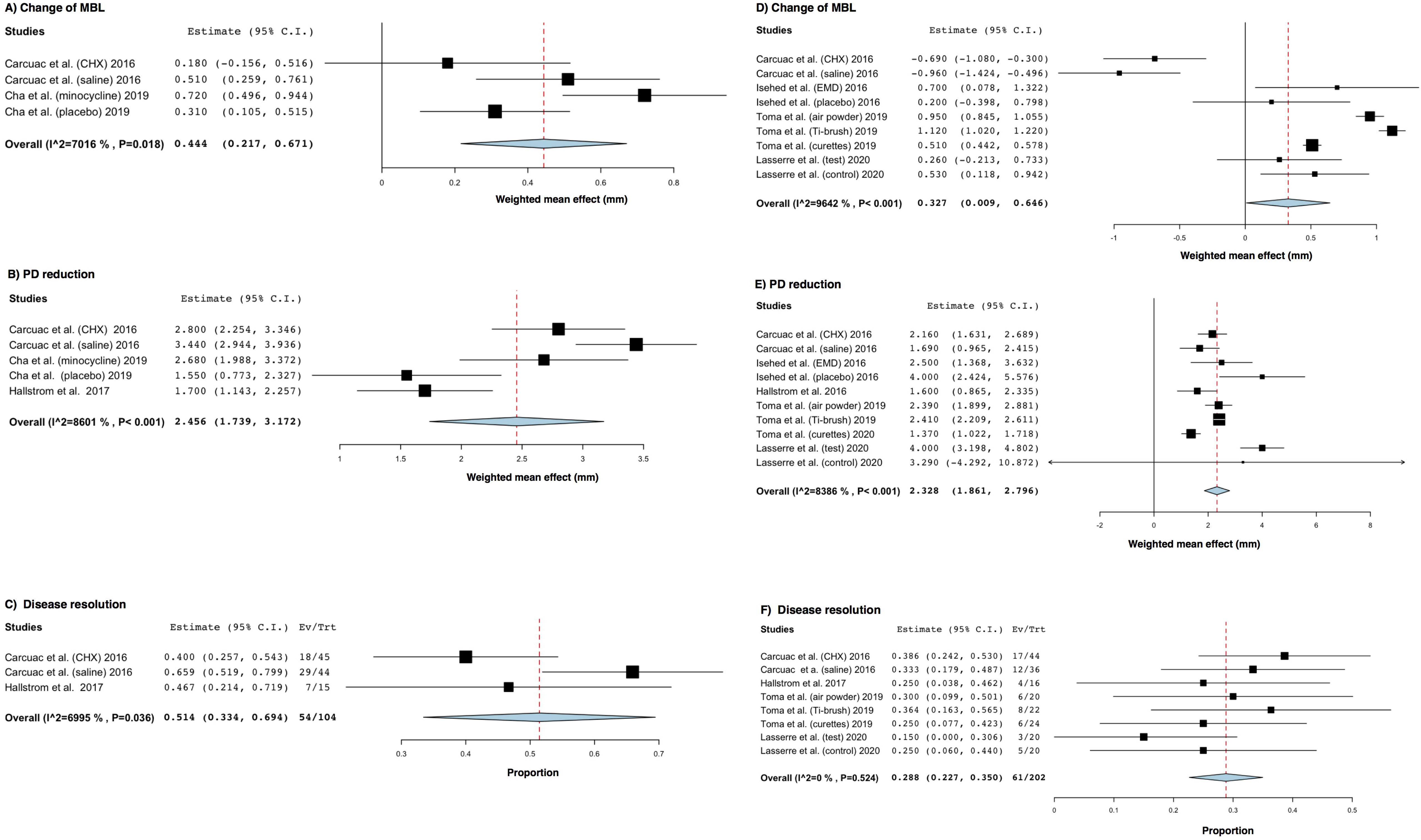
Forest plots of weighted mean effect (WME) for different clinical parameters after open flap debridement (OFD) with or without systemic antibiotics A) Marginal bone levels (MBL) changes with systemic antibiotics, B) Pocket depth (PD) reduction with systemic antibiotics, C) Disease resolution with systemic antibiotics. D) MBL changes without systemic antibiotics, E) PD reduction without systemic antibiotics, F) Disease resolution without systemic antibiotics.

Considering OFD without adjunctive systemic antimicrobials (Figures 4D,E,F) the WME of MBL amounted to 0.33 mm (95% CI: 0.01/0.65), based on nine arms of four studies with high heterogeneity. PD reduction was assessed in five studies (10 arms) and amounted to 2.33 mm (95% CI: 1.66/2.80). Finally, DR was calculated in three studies (five arms) with a percentage of 29% (95% CI: 23/35) and no heterogeneity.

ORS was performed with or without systemic antibiotics but no study provided a direct comparison, thus, it was not possible to perform a pair-wise meta-analysis. As depicted in Figure 5, only two arms of the same study reported data on MBL changes 12 months after ORS combined with systemic antibiotics. The WME was −0.27 mm (95% CI: −0.82/0.29) with significant heterogeneity.^45^ Four arms in two different studies from the same research group used ORS without systemic antibiotics.^34, 35^ WME for MBL accounted for −0.40 mm (95% CI: −0.67/0.13) with significant heterogeneity (*I*^2^= 81.76; p < 0.001), WME for PD reduction was 1.81 mm (95% CI: 1.59/2.03) with non-significant heterogeneity (*I*^2^= 23.05, p = 0.273). The same studies reported DR in a total of 26 successfully treated implants out of 171. This accounted for a weighted DR percentage of 11% (95% CI: 1/22) with significant heterogeneity (*I*^2^= 83; p < 0.001).

**Figure 5.**
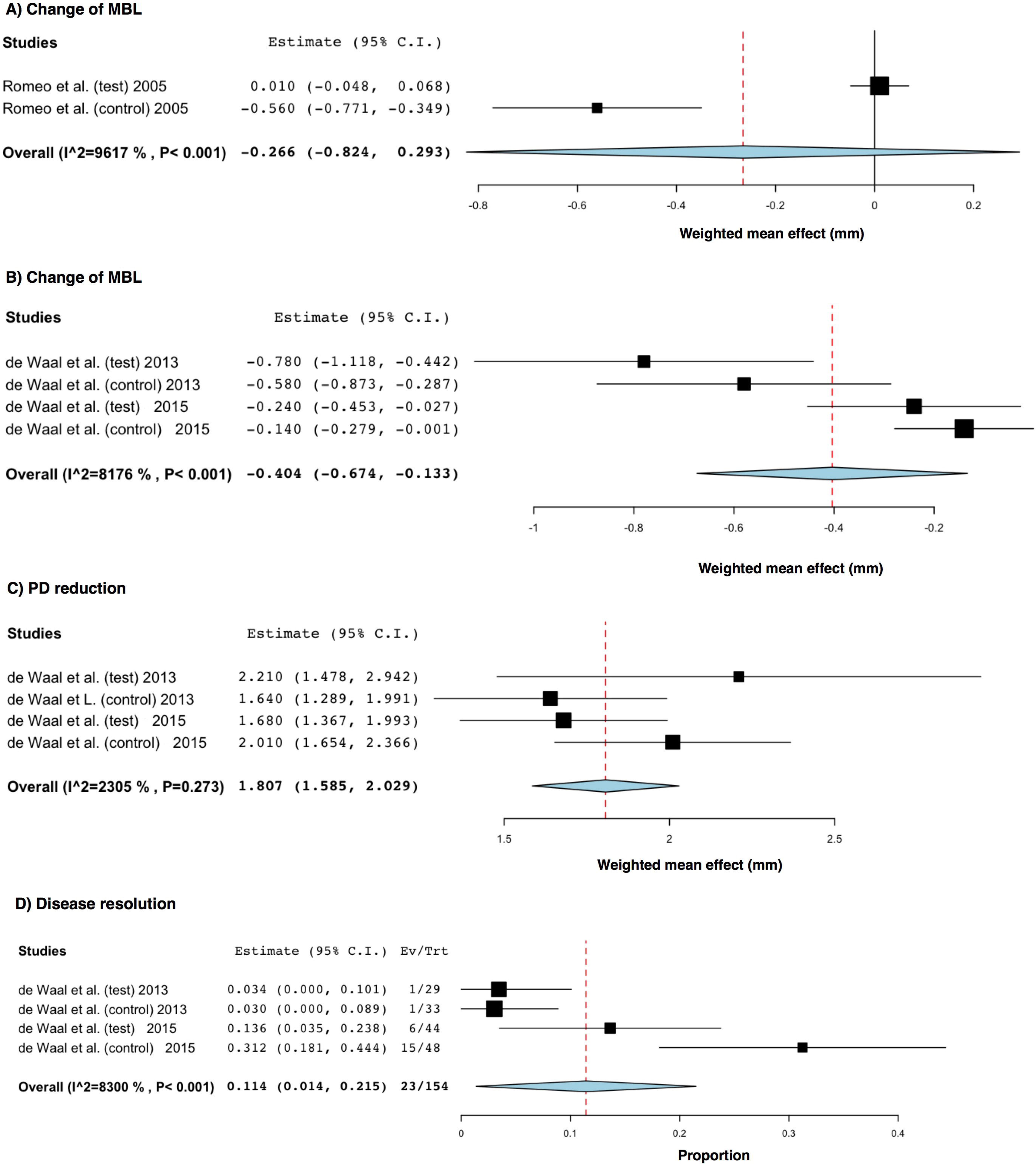
Forest plots of weighted mean effect (WME) for different clinical parameters after osseous resective surgery (ORS) with or without systemic antibiotic. A) Marginal bone levels (MBL) changes after ORS with systemic antibiotics, B) MBL changes after ORS without systemic antibiotics, C) Pocket depth (PD) reduction after ORS without systemic antibiotics, D) Disease resolution after ORS without systemic antibiotics.

## 4. Discussion

Peri-implantitis has become a rather prevalent complication with the increasing placement of dental implants for the rehabilitation of missing teeth.^47, 48^ Infection control has a pivotal role in the treatment of peri-implantitis, and there is a need for reliable and replicable methods. The present systematic review aimed at evaluating the efficacy of different anti-infective protocols for the surgical treatment of dental implants with clinical diagnosis of peri-implantitis. Overall, 16 RCTs were included in the qualitative and 9 in the quantitative analysis. Cohort studies or case series were excluded to obtain a higher quality of data and decrease the risk of bias.

### 4.1 Surface decontamination

Implant surface decontamination represents a fundamental step to achieve resolution of peri-implantitis lesions. However, the available clinical protocols show great variation and there is no evidence for the most effective procedure.^5^ Generally, there is a distinction between mechanical and chemical/physical decontamination methods, although a combination is usually recommended.^12^ Unfortunately, due to a significant heterogeneity within and among RCTs in terms of study design (definition of peri-implantitis, outcome, decontamination protocols, supportive care) and characteristics of the population (patient, implant and prosthetic features), a pairwise comparison between decontamination strategies was not possible and it remains unclear whether one protocol would provide greater benefits than others. However, the performances of different methods have still been assessed pooling data across studies.

OFD represents the best model for studies investigating clinical impact of different surface decontamination protocols at dental implants as it enables evaluation of the individual effect of the particular decontamination method on the final outcomes. Curettes are the most commonly used instruments for mechanical decontamination. Four study arms employed curettes alone to clean the implant surface during OFD. Improved PD values were reported (WME = 2.14 mm), while no significant differences were found in MBL changes (WME = −0.05 mm), and the effect in disease resolution was very modest (25%). It has to be kept in mind that the type of curettes (titanium or plastic) may account for a small variability in the findings.^49^ The RCT by Toma et al.^32^ offers a direct comparison between three different methods of mechanical decontamination. The titanium brush and glycine air-polishing devices were more effective than the plastic curettes, but treatment success remained low for all three treatment modalities (33%, 29% and 22%, respectively).

The two RCTs by de Waal et al.^34, 35^ indicated no additional significant benefit for the adjunctive employment of 0.12% CHX + 0.05% CPC with ORS. It is likely that implant surface decontamination may have less prominent effect when ORS is combined with apically positioned flap as a submucosal part becomes usually exposed by the surgical procedure. Moreover, there is controversial evidence on the role of implantoplasty. Romeo et al.^27, 45^ reported superior 3-year clinical and radiographic results with this intervention. On the contrary, no clinical benefit of implantoplasty on implant survival rate was observed in a long-term retrospective study, with other factors such as the initial defect depth and the frequency of recalls accounting for a larger influence on the outcomes.^50^ However, maintenance may be easier for smooth surfaces than surfaces with micro- and macro-plaque-retentive elements.

Ideally, implant surface decontamination should remove biofilm without causing surface damage not to render surfaces more conducive to bacterial colonization.^14^ *In vitro* studies indicated that non-metal curettes and rubber cups were minimally traumatic but ineffective to clean contaminated titanium surfaces; while ultrasonic scalers, metal curettes and rotating titanium brushes were effective particularly on modified titanium surfaces.^51^ The air abrasive system was effective in all types of implant surfaces, with glycine powder causing less alterations than sodium bicarbonate.^11, 52^ Nevertheless, *in vitro* studies do not properly simulate accessibility to the implant surface during surgical intervention. Anatomical factors and thread geometry can limit access of the decontamination devices, and implants with lower thread pitch and thread depth values may cause insufficient cleaning especially for instruments such as plastic curettes and air powder inserts with less flexibility.^53^

Removal of bacterial biofilm from contaminated implant surfaces with air-powder abrasion has been reported to provide significantly superior microbiological outcomes than chemical decontamination with hydrogen peroxide and chlorhexidine gluconate.^54^ Coupling mechanic instruments with chemical/physical agents may improve the overall cleaning ability as the chemical agent may reach niches inaccessible for the instruments.^12^ Numerous chemical decontaminants have been tested, although their efficacy has been difficult to demonstrate. Different concentrations of CHX showed limited efficacy both in *in vitro* models and in clinical trials, as it exerts cytotoxic activity.^55^ Carcuac et al.^9^ failed to find an influence of chemical decontamination on 1- and 3-year success using 0.2% CHX regardless of the implant surface and even in terms of microbiological modifications. The present finding of lack of efficacy with 0.12% CHX + 0.05% CPC in ORS partially confirms the report of Carcuac et al.^9^

Review articles and controlled clinical studies indicated that local delivery of antibiotics combined with non-surgical treatment of peri-implantitis lesions improved clinical parameters.^56–58^ Particularly, repeated application of locally delivered minocycline during post-surgical follow-up enhanced the clinical and microbiological effect of OFD, achieving 0.72 mm of vertical bone gain and 66.7% of disease resolution.^36^

On the other hand, current evidence shows that laser or photodynamic therapy in combination with surgical/non-surgical treatment of peri-implant diseases provided minimal benefit in PD reduction, CAL gain, and MBL improvements.^59, 60^ However, properly controlled clinical trials with low risk of bias are warranted to better clarify the issue.

Within the context of surgical regenerative treatment of peri-implantitis, several decontamination techniques have been described, but the large heterogeneity in flap design, biomaterials, bone defect anatomy and post-operative care restricts the conclusion on their efficacy. Encouraging outcomes were obtained by Isehed et al.,^46^ with EMD applied on fixture surfaces switching subgingival microbiota to Gram+ aerobic populations. This ecological shift was linked with an increase in bone levels as compared with non-EMD controls. Decontamination of the supra-and the intra-osseous part of the defect-adjacent implant surface has a particular importance. To achieve this, Schwarz et al.,^41^ proposed a combined approach with regenerative treatment attempted apically and a resective approach with implantoplasty carried out in the supra-osseous part of the fixture, where re-osseointegration cannot be achieved. The adoption of the combined approach yielded a 66.7% of disease resolution in a RCT which used titanium brushes to decontaminate the intra-osseous part of the implant.^43^

### 4.2 Adjunctive systemic antimicrobials

Systemic antimicrobials are likely to have a place in the anti-infective strategies for implant surface decontamination and they have been traditionally prescribed in spite of the debate on global concerns of bacterial resistance development.^61^ Meta-analyses identified significantly larger MBL gain (WMD = 1.17 mm) with peri-surgical systemic antimicrobials, whereas probability of disease resolution (RR = 1.50) and differences in PD reduction approached statistical significance. Moderate to substantial heterogeneity was found and none of the included studies addressed PROMs. Carcuac et al.^21^ employed four parallel study arms to evaluate the single and combined effect of local decontaminant and systemic antimicrobials in OFD. While no added benefit was observed for CHX, a significant adjunctive effect in terms of MBL, PD reduction and DR was reported for the two groups with systemic antimicrobial prescriptions. Conversely, Hallström et al.^17^ did not find significant differences with systemic antibiotics. This discrepancy could be partially explained by different sample sizes or type and regimen of antibiotics used in these two different RCTs.^62^ The systemic antibiotic was started three days before the surgery and continued for 10 days in the RCT by Carcuac et al.,^21^, emphasizing the importance of pre-operative infection control. Overall, the findings of the present systematic review support the use of systemic antibiotics in the surgical treatment of peri-implantitis, even though a careful risk/benefit analysis is mandatory considering PROMs and the growing issue of antibiotic resistances. The available RCTs included in this review fail to adequately address these aspects.

### 4.3 Limitations and suggested future research

The present study has some limitations mainly related with the nature of the available literature. Several factors such as implant surface characteristics,^63^ configuration of peri-implant defect,^64, 65^ frequency and quality of supportive therapy^8, 66^ can affect the results of surgical treatment of peri-implantitis, but none of these factors could be considered in the present meta-analyses. Furthermore, 6-month follow-up in some of the included RCTs may be too short for detection of significant effect on MBL changes and may not account for the relatively high incidence of disease recurrence following surgical therapy of peri-implantitis.^67^ This threshold was chosen for two reasons: firstly to increase the number of eligible studies; and secondly to focus on the short-term clinical outcomes that are more likely to be explained by the decontamination procedures.

To the best of our knowledge, this systematic review is the first attempting to provide a qualitative and quantitative evaluation of the clinical outcomes of anti-infective protocols across different surgical procedures reported in published RCTs. This systematic review had a rather wide scope including RCTs using regenerative, resective, and combined surgery. This approach and the information outlined in Table 2 may provide valuable insight for the clinicians in treatment planning. The 8th European Workshop on Periodontology stated that “a proven method of decontaminating the implant surface” is a critical component of surgical treatment, yet available clinical, radiographic, and microbiological data do not favor any decontamination approach in surgical therapy so far.^68^ However, this finding does not mean that no treatment option is effective,^69^ but there is just no consensus on the best treatment procedure. Reasons may be imputable to different advantages and disadvantages associated with each method, and the tendency to adopt multiple combinations of decontaminants to increase the likelihood of success. Well-designed *in vitro* and *in vivo* studies with adequate sample size allowing comparisons with low risk of bias are warranted in order to establish an evidence-based protocol for decontamination during the surgical treatment of peri-implantitis.

## 5. Conclusion

There is lack of evidence to support one intra-surgical protocol of implant surface decontamination over others. The use of curettes during OFD is able to produce PD reduction but has limited effect on MBL and DR; while different CHX formulations do not offer any benefits. Lack of replication was observed for all other chemical and mechanical decontamination methods and future research should aim for higher consistency. In parallel, the use of adjunctive systemic antimicrobials with the surgical treatment of peri-implantitis seems to be supported by a tendency for a clinical improvement at short-term, even though the risk/benefit ratio should be cautiously assessed by the clinicians.

## Data Availability

All data generated or analyzed during this study are included in this published article [and its supplementary information files].

---

**Supplementary Figure 1.** PRISMA flow diagram depicting the selection process.

## Notes

**Conflict of Interest Statement** The authors declare no conflict of interest.

**Availability of data and material** All data generated or analyzed during this study are included in this published article [and its supplementary information files].

### Competing Interest Statement

The authors have declared no competing interest.

### Clinical Trial

PROSPERO CRD42020182303

### Clinical Protocols

https://www.crd.york.ac.uk/prospero/display_record.php?ID=CRD42020182303

### Funding Statement

This study was funded solely by the institutions of the authors.

### Author Declarations

This article does not contain any studies with human participants performed by any of the authors.

